# Access to malaria diagnosis and treatment in Zambia in the context of scaling-up community case management: results from repeated national cross-sectional surveys

**DOI:** 10.64898/2026.08.25.26361362

**Authors:** Zhiyuan Mao, Adam Bennett, Kafula Silumbe, John Miller, Justin Millar, Hannah Slater, Josh Yukich, Ruth A. Ashton, Irene Kyomuhangi, Andrew Andrada, Refilwe Karabo, Thomas P. Eisele

## Abstract

**Background:** Community case management has been scaled up nationally in Zambia over the last decade. However, there is limited evidence on how this nationwide implementation has affected febrile patients’ access to malaria diagnosis and treatment in Zambia.

**Methodology:** This study analyzed four rounds of Malaria Indicator Survey (MIS) data (2012-2021) to evaluate: 1) proportion of all-ages individuals with fever who sought treatment from a formal provider, 2) proportion of individuals going to CHWs over time, among those who sought treatment at a formal provider, 3) time duration between fever onset and treatment seeking at a formal provider, and 4) proportion of children <5 with malaria that received Artemether-Lumefantrine (AL) treatment. Mixed-effect logit models were employed to examine determinants of treatment-seeking behavior and factors affecting AL receipt among children <5 with malaria cases.

**Results:** The proportion of febrile patients seeking treatment remained below 60% throughout 2012-2021, and AL receipt among children with malaria cases consistently stayed below 50%. The mean interval between fever onset and initial treatment-seeking encounter decreased from 2.42 days in 2012 to 1.71 days in 2021. Among formal care seekers, CHW utilization increased from 1.5% in 2012 to 10.0% in 2018 before declining to 3.2% in 2021. Longer walking time to the nearest health facility was associated with lower odds of treatment seeking, whereas CHW density was not associated with treatment seeking or AL receipt. Children who did not went to formal providers had lower odds of AL receipt than those who sought treatment from CHWs.

**Conclusion:** Despite nationwide CCM scale-up over the last decade, significant barriers persist in malaria patients’ access to diagnosis and treatment in Zambia. Our results indicate that while CCM coverage should be maintained and further expanded, additional complementary interventions are also needed to overcome remaining access barriers.

## Introduction

Malaria, primarily caused by *Plasmodium falciparum*, poses a serious public health threat in Zambia. In 2022, it was estimated there were over 3.5 million malaria cases and over 8,500 malaria deaths.(1) Despite the high number of malaria cases, Zambia has made significant strides in combating malaria over the past 20 years.(2–5) Over this period, Zambia has made notable progress in scaling up vector control and strengthening malaria diagnosis and treatment. It was the first country in Africa to adopt artemether-lumefantrine (AL) as the first line treatment for uncomplicated malaria in 2002.(4) Since 2006, rapid diagnostic tests (RDTs) have been integrated into parasitological examination in health facilities, as well as surveillance efforts.(5)

Ensuring access to prompt malaria diagnosis and treatment remains a significant challenge among remote rural populations with limited access to health care, where the risk of malaria is greatest.(6) Although Zambia has committed to universal health care, substantial geographic, infrastructural, and health spending constraints continue to limit effective service delivery and outreach in many rural and hard-to-reach areas.(7,8) Community case management (CCM) has emerged as a key solution to this problem.(9) Through CCM, community health workers (CHWs) are equipped to provide parasitological testing using RDTs, administer the first line drug AL for confirmed malaria cases, and facilitate referrals to higher level care for patients exhibiting severe malaria symptoms.(10) Previous studies have linked the expansion of CCM to improved coverage of vector control measures, increased use of intermittent preventive treatment in pregnancy (IPTp), as well as improved access to prompt malaria diagnosis and treatment.(6,11,12)

Zambia’s Malaria Elimination Strategic Plan 2022-2026 has emphasized universal access to early diagnosis and treatment within 24 hours of fever onset.(13) This approach is facilitated through public facilities and CCM, as well as the private sector in some contexts. In line with this, the National Community Health Strategy (NCHS) 2019-2021 set a goal of ensuring that all Zambians can access quality basic health services within 1 hour of travel distance by 2021, primarily relying on the expansion of CCM.(14) Starting from 2013, CHWs in Zambia have been trained to deliver outreach services, addressing the shortage of doctors and nurses.(15) These services include promoting personal hygiene practices and providing basic healthcare, particularly in malaria diagnosis and treatment, as well as management of HIV/AIDS in some settings.(16) The first pilot study of CCM for malaria, launched by the Zambia National Malaria Elimination Centre (NMEC), occurred in Lusaka and Southern Province in 2009.(17) Following that, CCM was scaled-up in an incremental fashion at national level between 2013-2020.(15) Research by Ashton and colleagues (2023) showed there to be an association between the CCM expansion and a reduction in inpatient malaria cases and deaths across Zambia.(15) However, except for this research, there is limited evidence showing the impact of CCM expansion on access to malaria diagnosis and treatment in Zambia. This study aimed to examine the association between CCM expansion and access to malaria diagnosis and treatment, and to identify determinants of treatment-seeking behavior among febrile individuals across all ages, as well as factors associated with a child with malaria receiving AL.

## Methodology

### Ethics statement

This study utilized data exclusively from the Zambia Malaria Indicator Surveys (MIS), conducting secondary analysis of these de-identified datasets. No primary data collection was performed. The datasets were accessed for research purposes on [06/01/2023]. The authors did not have access to any information that could identify individual participants during or after data collection. In accordance with standard research practices for secondary analyses of anonymous public health surveillance data, formal ethics approval was not required for this study.

#### Data

We used individual-level data from the nation-wide Zambia MIS conducted in the peak malaria transmission months of April and May for the years 2012, 2015, 2018, and 2021. Each survey employed a two-stage probability proportional to size (PPS) cluster sampling design, with the number of standard enumeration areas sampled ranging from 113 to 202. Variables in the MIS dataset used in our analyses included histidine rich protein 2 (HRP2) RDT results among children under 5 years old, using First Response® Malaria AG HRP2 in 2012 and Bioline® P.f HRP2 RDT in 2015, 2018, and 2021, fever history in the past two weeks, treatment providers for fever, age, sex, household head’s educational attainment, household socioeconomic status (SES), urbanicity, province, survey cluster, and household GPS coordinates.

We integrated data from various sources into the MIS dataset linked to time and space. Information about CHWs and operational health facilities for each survey period were sourced from the District Health Information Software 2 (DHIS 2). Instead of calculating the straight-line distance (Euclidean distance), We used a ‘Walking-only friction surface’ (1km x 1km resolution) to estimate foot-based travel time from individual household to malaria providers.(18) This approach was used because household-level data on access to motorized or non-motorized transportation were not available, and walking represents the most consistently observable and comparable mode of travel across settings. Local *P. falciparum* parasite rate among children aged 2-10 (PfPR_2-10_) was obtained from the Malaria Atlas Project (MAP).(19) Average precipitation over a 5-day span, from Climate Hazards Group InfraRed Precipitation with Station Data (CHIRPS Pentad),(20) was integrated into our study to examine the potential impact on the association between CCM scale-up and treatment seeking behavior change. Household SES of MIS respondents was determined by applying principal component analysis to a list of household assets collected during the survey. Additionally, we included geo-referenced population density from World Pop 3 for the years 2012, 2015, 2018, and 2021 (matched to corresponding MIS survey years),(21) and the geo-referenced relative deprivation index from the Center for International Earth Science Information Network (CIESIN) at Columbia University.(22) This index is a composite measure that reflects relative socioeconomic deprivation at the community level, incorporating indicators such as access to infrastructure, housing quality, and basic services. The relative deprivation index was used as a proxy for the scarcity of key community resources in each household location.

#### Study population and treatment providers for fever

The populations used in this study were febrile individuals of all ages, and children <5 years old with a current or recent malaria case at the time of each MIS visit. We defined a febrile individual as anyone reporting fever symptoms within 14 days prior to the survey visit. A recent or current malaria case was defined as children <5 years old who met the following criteria: a) reported a fever within 14 days prior to the survey visit; and b) had a positive HRP2 RDT at the time of the survey visit.(23) This approach is supported by the fact that a positive RDT indicates current/recent exposure to *P. falciparum*, as the HRP2 antigen remains detectable even after previous AL treatment up to 2-3 weeks, with this detection period overlapping with the patient’s history of fever for the past two weeks.(24–26)

MIS respondents who stated they had a fever in the previous two weeks and sought treatment for the fever were asked where they sought treatment, based on 12 pre-defined healthcare categories. We regrouped these responses into three formal provider categories: public providers, CHWs, and private providers. “Government Hospital,” “Government Health Center,” “Mobile Clinic,” and “Health Post” were classified as public providers. CHW care was restricted to respondents who explicitly reported seeking care from a “Field Worker.” “Private Hospital,” “Private Clinic,” and “Private Doctor” were classified as private providers. Responses such as “Local Shop,” “Traditional Practitioner,” and “Pharmacy” were classified as not seeking care from a formal provider. These responses are recorded under an ‘Other’ category in the MIS and are inconsistently specified, limiting reliable classification of provider type or diagnostic capacity. Moreover, these outlets do not consistently provide quality-assured malaria diagnosis and treatment. Additionally, we considered instances where febrile individuals visited multiple providers. However, reported treatment-seeking from more than one type of health facility was rare, with less than 1% of individuals reporting this in 2012, 2018, and 2021, respectively. For those who sought treatment from both a CHW and a public/private provider, we assumed they were referred by the CHW, and thus these cases were exclusively categorized under CHW.

#### Primary and secondary outcomes

The primary outcomes of our study were indicators of enhanced access to malaria diagnosis and treatment in Zambia: 1) proportion of all-ages individuals with fever who sought treatment from a formal provider, 2) proportion of individuals going to CHWs over time, among those who sought treatment at a formal provider, 3) time duration between fever onset and treatment seeking at a formal provider, and 4) proportion of children <5 with malaria that received AL treatment. There were also two secondary outcomes: 1) determinants of treatment-seeking behavior from formal providers; and 2) factors affecting AL receipt among children <5 with malaria.

#### Travel time to nearest provider

Our analysis estimated each MIS participant’s walking time to the nearest formal provider. We hypothesized that in the Zambian context, where many rural communities have limited transportation options and challenging terrain, walking time would be the most significant driver of treatment-seeking behavior among febrile patients.

We used the Walking-only Friction Surface together with household GPS coordinates from MIS and location data of formal provider (either CHW or health facility) from DHIS2 to calculate travel times. Active CHWs were defined as those reporting at least one malaria case between April 1 and June 1 of each survey year, which aligns with the high transmission season and MIS fieldwork timing. CHWs were identified through the malaria case reporting system using CHW identifiers; a comprehensive master list of all CHWs in Zambia was not available. Accordingly, this definition was intentionally restricted to CHWs engaged in malaria diagnosis and treatment activities, as the analysis focused on access to malaria diagnosis and treatment. CHWs engaged in other health activities but not reporting malaria cases during this period would therefore not be classified as active in this context. Only facilities with complete geographic location information were included in the analyses. We assumed that health facilities recorded in DHIS2 with valid location data remained operational during the corresponding survey periods. Consequently, for 2012 (pre-CCM expansion), our analysis only included health facilities as providers. For subsequent survey years (2015, 2018, and 2021), the analysis incorporated both health facilities and active CHWs. All spatial coordinates were projected using Arc 1950 / UTM zone 36S (EPSG:20936) to calculate travel times from each respondent to their nearest formal provider. This approach accounts for natural barriers and road conditions that simple straight-line distances would not capture. For survey households without valid GPS coordinates, the centroid of the reported district was used as a proxy location for travel-time estimation.

#### CCM expansion

Our study, furthermore, quantified CCM expansion by calculating the ratio of CHWs per capita at the district level for each of the four survey years. This measurement was done by identifying and counting the number of CHWs in each district. Provider locations were spatially matched with district boundaries. For each district and survey year, we derived the ratio of providers to population, presenting results as the number of providers per 1,000 population. In addition, these district-level ratios were then aggregated to the provincial level, adjusting for the relative population size of each district to ensure appropriately weighted provincial estimates.

#### Statistical analyses

##### Descriptive statistics

To assess whether the CCM expansion enhanced access to malaria diagnosis and treatment in Zambia from 2012 to 2021, we estimated each primary outcome with four rounds of MIS (2012-2021). Each survey round was analyzed independently, without pooling the data across years, and the estimates were adjusted for sampling weights. For stratified analyses, such as the overall treatment-seeking rate by province and gender, data from the four survey rounds were pooled, with the weights recalculated accordingly. In addition, we constructed a panel of province–year scatter plots relating CHW density to the primary outcomes to descriptively assess heterogeneity across levels of CCM expansion. These correlations are unadjusted, pooled across survey years, and do not represent causal or within-year effects.

#### Analyses of associations

Our research first applied mixed-effects logistic regression models to investigate the determinants of treatment-seeking behavior in febrile individuals of all ages (Model 1). For this model, in addition to travel time to the nearest HF (in hours) and number of CHW per 1,000 population at district level, covariates including age group (categorized as 0-4, 5-14, and 14+ years), urbanicity, survey year, precipitation, *PfPR*_2-10_, population density, relative deprivation index, quintile household SES, household head’s education attainment and province were modelled as the fixed-effect variables, while survey cluster was treated as the random intercepts. To address the non-linear relationship between travel time and treatment-seeking behavior, a squared term for travel time was included in the model. Additionally, the potential interaction between travel time and age group was tested in the model. The final model was presented in the manuscript.

For the analysis of AL receipt in children <5 years with malaria (Model 2), a mixed-effects logit model was also implemented. For this model, apart from travel time to the nearest HF (in hours) and number of CHW per 1,000 population at district level, the covariates included formal provider, age in years, gender, survey year, *PfPR*_2-10_, household head’s education attainment, household SES and province. Survey clusters were treated as random intercepts to account for intra-group correlation.

All analyses were conducted in the R programming environment (R Version 4.3.1) with models developed by glmmTMB, buildmer, and see packages.(27–30)

## Results

A total of 9,913 febrile individuals out of 69,984 surveyed participants (14.2%) were included in the analysis of treatment-seeking behavior in Zambia across four rounds of Malaria Indicator Surveys (MIS 2012: 16,928 participants; MIS 2015: 16,145 participants; MIS 2018: 18,384 participants; MIS 2021: 18,527 participants). Among these febrile individuals, 1,394 children under the age of 5 with recent or current malaria cases were specifically investigated to determine the receipt of AL.

From 2012 to 2021, the overall proportion of individuals seeking treatment for fever in the previous 2 weeks remained above 55.0% across the Zambia MIS’s, with no significant differences between survey years (Table 1). Overall treatment seeking also did not differ significantly by age, sex, household SES, urbanicity, or educational attainment of the household head (supplementary material Table 1).

**Table 1.** Trends in malaria treatment-seeking behaviors and outcomes in Zambia, 2012-2021 Malaria Indicator Surveys.

| Metric | Estimates |
| --- | --- |
| Proportion of all-ages individuals with fever who sought treatment from a formal provider |  |
| 2012 | 55.8% [52.0%, 59.5%] |
| 2015 | 56.0% [52.5%, 59.4%] |
| 2018 | 59.0% [53.5%, 64.4%] |
| 2021 | 57.1% [52.4%, 61.7%] |
| Proportion of individuals going to CHWs, among those who sought treatment at a formal provider |  |
| 2012 | 1.5 % [0.7%, 2.7%] |
| 2015 | 7.6% [4.6%, 11.7%] |
| 2018 | 10.0% [4.8%, 17.9%] |
| 2021 | 3.2% [1.2%, 6.8%] |
| Time duration between fever onset and treatment seeking at a formal provider |  |
| 2012 | 2.42 days [2.27, 2.58] |
| 2015 | 2.00 days [1.87, 2.14] |
| 2018 | 2.39 days [2.08, 2.70] |
| 2021 | 1.71 days [1.54, 1.89] |
| proportion of children <5 with malaria that received AL treatment |  |
| 2012 | 38.4% [31.3%, 45.8%] |
| 2015 | 40.7% [34.0%, 47.6%] |
| 2018 | 45.0% [36.0%, 54.3%] |
| 2021 | 41.7% [34.2%, 49.5%] |

Of the febrile individuals of all ages who sought treatment at formal providers, the proportion who reported seeking care from an explicitly identified CHW increased from 1.5% in 2012 to 7.6% in 2015 and 10.0% in 2018, before declining to 3.2% in 2021 (Table 1). Public providers remained the most commonly reported source of care across all surveys. Since 2015, private providers were reported less frequently than CHWs (Table 2). When extending the analysis to include all febrile individuals—including those who did not seek treatment and those who sought from informal providers—we observed a similar pattern (Supplementary Table 3).

**Table 2.** Number and proportion of febrile individuals by type of formal healthcare provider in Zambia, 2012-2021 *Individuals who didn’t seek treatment from aformal provider were excluded.

| MIS year | Public facility | CHW | Private facility | Total |
| --- | --- | --- | --- | --- |
| 2012 | 1,138 (92.8%) | 27 (1.5%) | 50 (5.8%) | 1,215 |
| 2015 | 1,029 (89.2%) | 90 (7.6%) | 25 (3.2%) | 1,144 |
| 2018 | 1,212 (89.8%) | 120 (10.0%) | 3 (0.1%) | 1,335 |
| 2021 | 1,104 (95.7%) | 50 (3.2%) | 10 (1.2%) | 1,164 |

Among all febrile individuals seeking treatment, the duration between the onset of fever and their first healthcare encounter varied across survey years. The average duration in 2021 was significantly shorter (1.71 [1.54, 1.89] days) compared to the previous survey periods (2.42 [2.27, 2.58]; 2.00 [1.88, 2.14], and 2.39 [2.08, 2.70] days for 2012, 2015, and 2018, respectively, Table 1). The proportion of febrile individuals who sought treatment within 24 hours of fever onset also increased significantly in 2021 (45.3%) compared to 2012 (30.6%) and 2018 (23.0%).

Regarding receipt of AL among children <5 with malaria at the time of the survey, the proportion that received AL remained consistently below 50% from 2012 to 2021, with a slight, non-significant increase from 38.4% in 2012 to 41.7% in 2021 (Table 1). Similar to the trends in treatment-seeking behavior, no significant differences were found in the proportion of children with recent/current malaria cases that received AL by urbanicity and age groups (Supplementary material table 2). The ratio of malaria care providers per capita showed substantial growth from 2012 to 2021, with the national average increasing threefold from 0.31 to 0.93 providers per 1,000 population (Figure 1). However, this expansion varied markedly across provinces. Southern Province exceeded 1 provider per 1,000 population beginning in 2015, while Western Province rose above this threshold later (2018) but ultimately reached the highest density of 1.73 [1.01, 2.36] by 2021. Northwestern and Eastern Provinces demonstrated marked improvements primarily in the later period, increasing from baseline ratios of 0.4 to 1.31 [0.54, 2.02] and 1.19 [0.12, 2.11] respectively by 2021. The remaining provinces (Lusaka, Luapula, Northern, Muchinga, Central, and Copperbelt) exhibited modest and uneven growth, with increases concentrated in a subset of districts, while many districts showed little or no change; final provider ratios ranged from 0.22 to 0.70 per 1,000 population.

**Figure 1.**
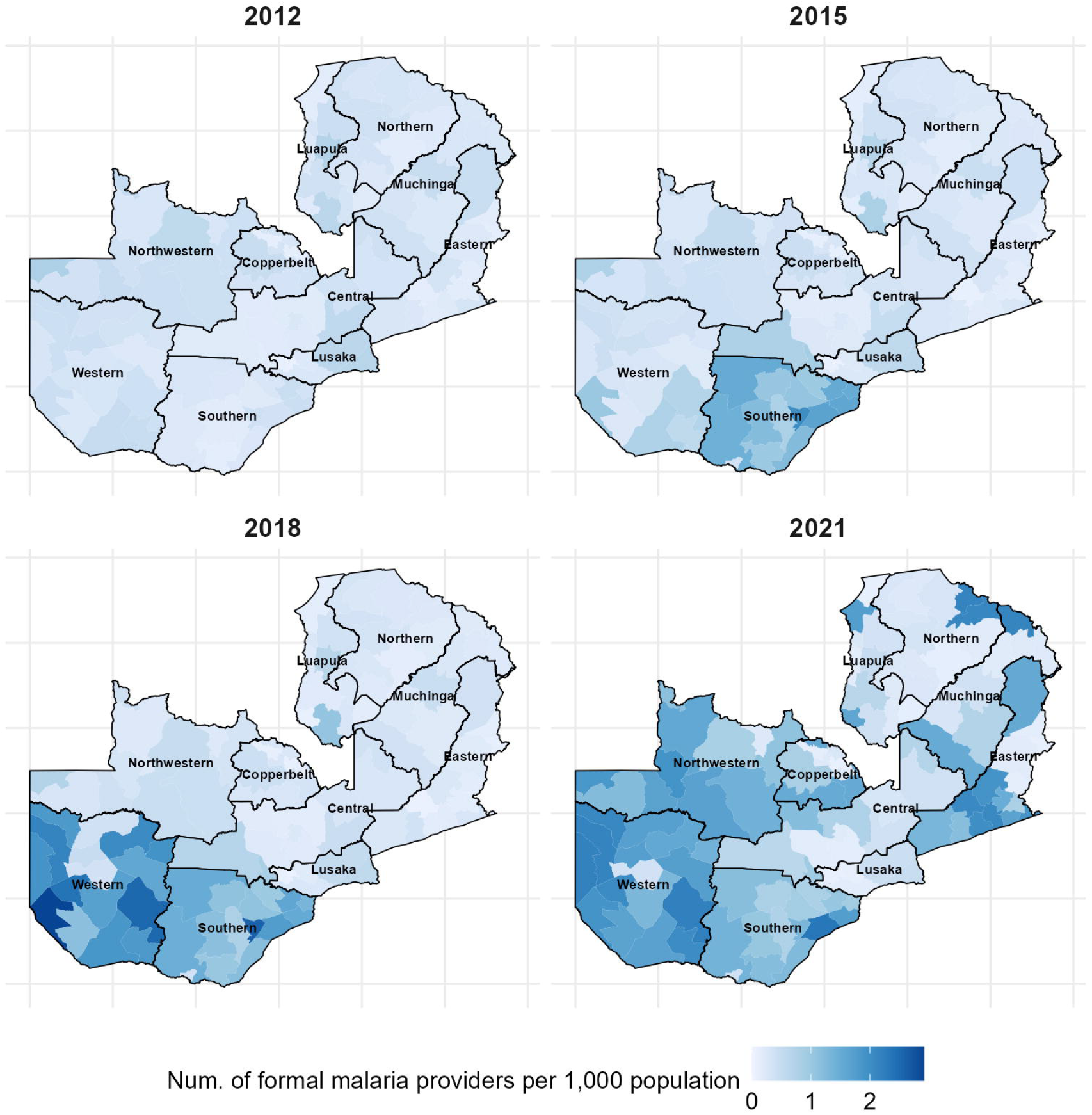
Distribution of malaria care providers per capita across Zambia, 2012-2021. * Formal malaria care providers include both health facilities and CHWs. The analysis assumes all health facilities remained continuously operational throughout the study period (2012-2021), while the number of CHWs varied annually across provinces. Base map source: GRID3, “ZMB – Operational Districts”dataset (https://data.grid3.org/datasets/zmb-operational-districts/explore), licensed under CC BY 4.0.

Provincial estimates for primary outcome and walking time to nearest provider (including both CHWs and public facilities) are presented in Supplementary Figures S1–S5. Except for walking time, no consistent temporal patterns were evident across provinces. Nationally, the estimated mean walking time decreased from 41.9 minutes [34.3, 49.5] in 2012 to 30.4 minutes [25.5, 35.3] in 2021, with substantial improvements observed in South and West parts of the country. AL receipt estimates were unavailable for Southern and Lusaka provinces in several survey years because no children in those province–year samples met our definition of a recent or current malaria case. In descriptive province– year analyses excluding 2012, CHW density showed weak pooled correlations with treatment seeking (r=0.06), CHW share among formal seekers (r=−0.07), days to first care (r=0.03), and AL receipt (r=−0.02) (Fig 2).

**Figure 2.**
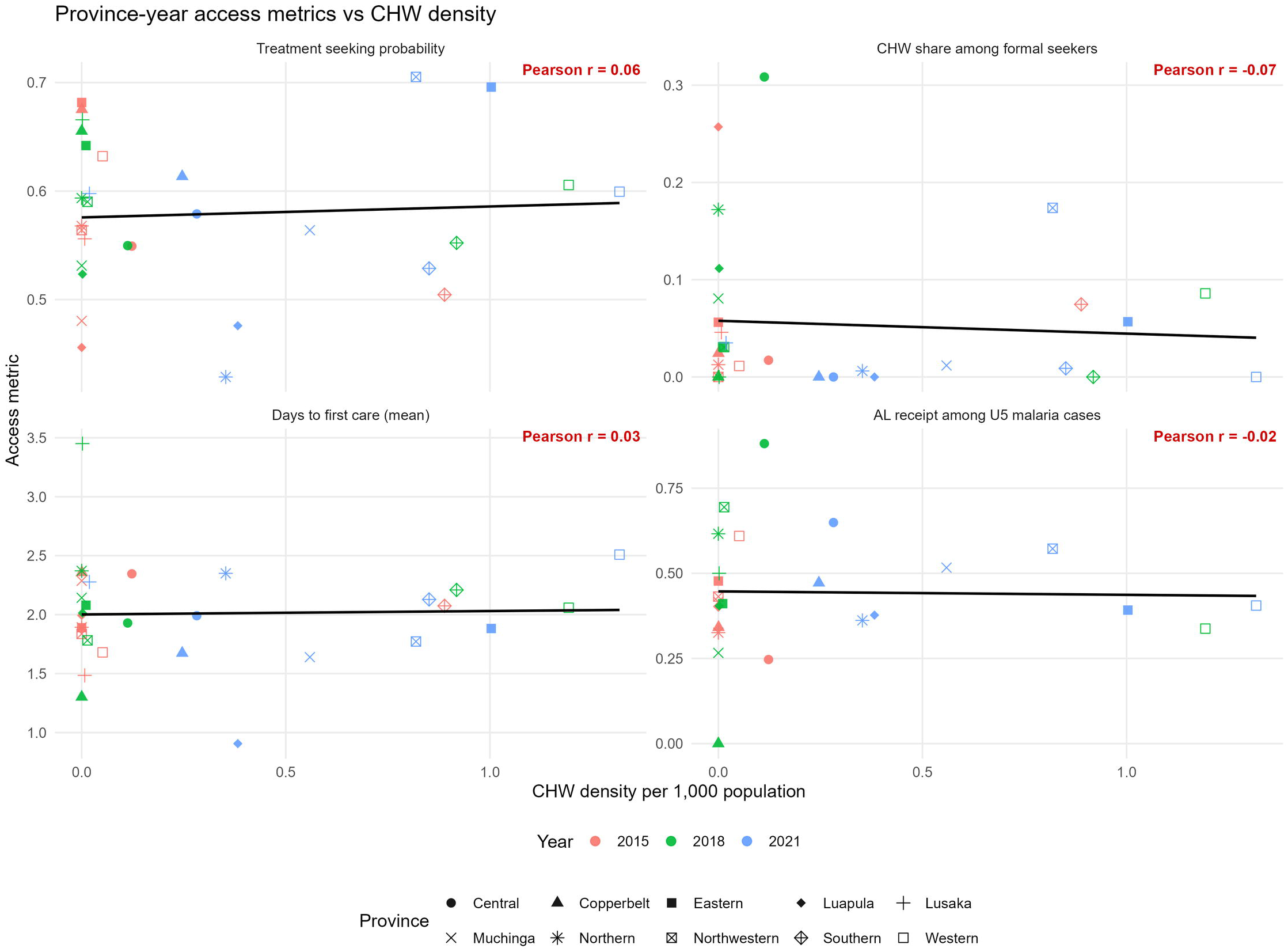
Association between CHW density and treatment seeking, CHW share among formal care providers, time to first medical encounter, and AL receipt among children with malaria. *Note:* Each point represents a province–year estimate. Data from 2012 were excluded because CCM expansion had not yet been implemented.

### Determinants of treatment-seeking behavior among febrile individuals (Model 1)

Across the survey rounds, older individuals were observed to be less likely than children <5 years old to have sought treatment for their fever (adjusted odds ratios (AOR) were 0.82 [0.77, 0.88] and 0.67 [0.61, 0.75] for the 5-14 and 14+ age groups, respectively, compared to the under-five age group). Household SES also impacted the treatment-seeking for fever, with high and highest SES households being more likely to do so compared to the lowest SES households (Figure 3). Longer traveling time to the nearest health facility was associated with lower odds of treatment seeking (AOR: 0.67 [0.56, 0.80]), with evidence of nonlinearity (1.07, [1.02, 1.12]). Also, CHW density (per 1,000 population) in the patient’s district of residence was not found associated with the likelihood of seeking treatment for fever. By controlling for other covariates, a provincial variation was still noticed. The interaction term between travel time and age group was found to be statistically non-significant and was therefore not included in the final model.

**Figure 3.**
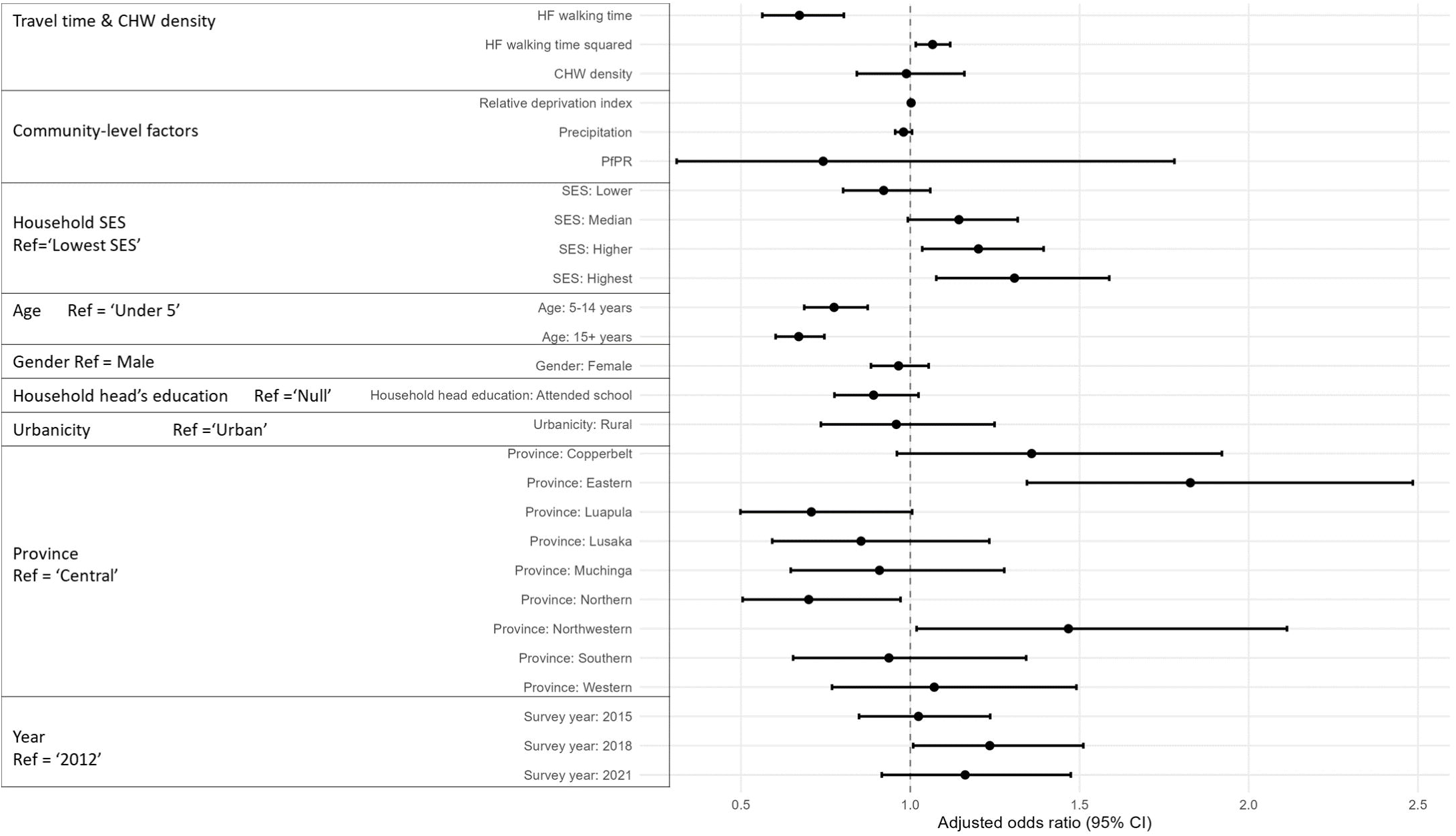
Model 1 outputs. The determinants of treatment-seeking behavior among febrile individuals. Results presented are the associated tio (OR), 95% confidence interval (CI) from the logit model. N=9,491. Note: A total of 422 observations with missing data on relative on index, household-head educational attainment, or PfPR were excluded from the complete-case analysis. Travel time was d in hours.

### Factors affecting AL-receipt among children <5 years old with malaria (Model 2)

Compared to children who sought treatment from CHWs, those who did not obtain care through a formal provider were substantially less likely to have received AL for their illness (AOR: 0.075 [0.03, 0.17]; Figure 4). The odds of AL receipt did not differ significantly between children who sought care from public or private providers and those who sought care from CHWs. The odds of AL receipt were similar across most provinces relative to Central Province. The exception was Southern Province, where children had significantly lower odds of receiving AL (AOR: 0.07 [0.01, 0.41]). No consistent socioeconomic gradient in AL receipt was observed, although the middle SES category differed significantly from the reference category. Of note, the travel time to nearest health facility, CHW density, survey year, and educational attainment of household heads were not significantly associated AL-receipt in this model (Figure 4).

**Figure 4.**
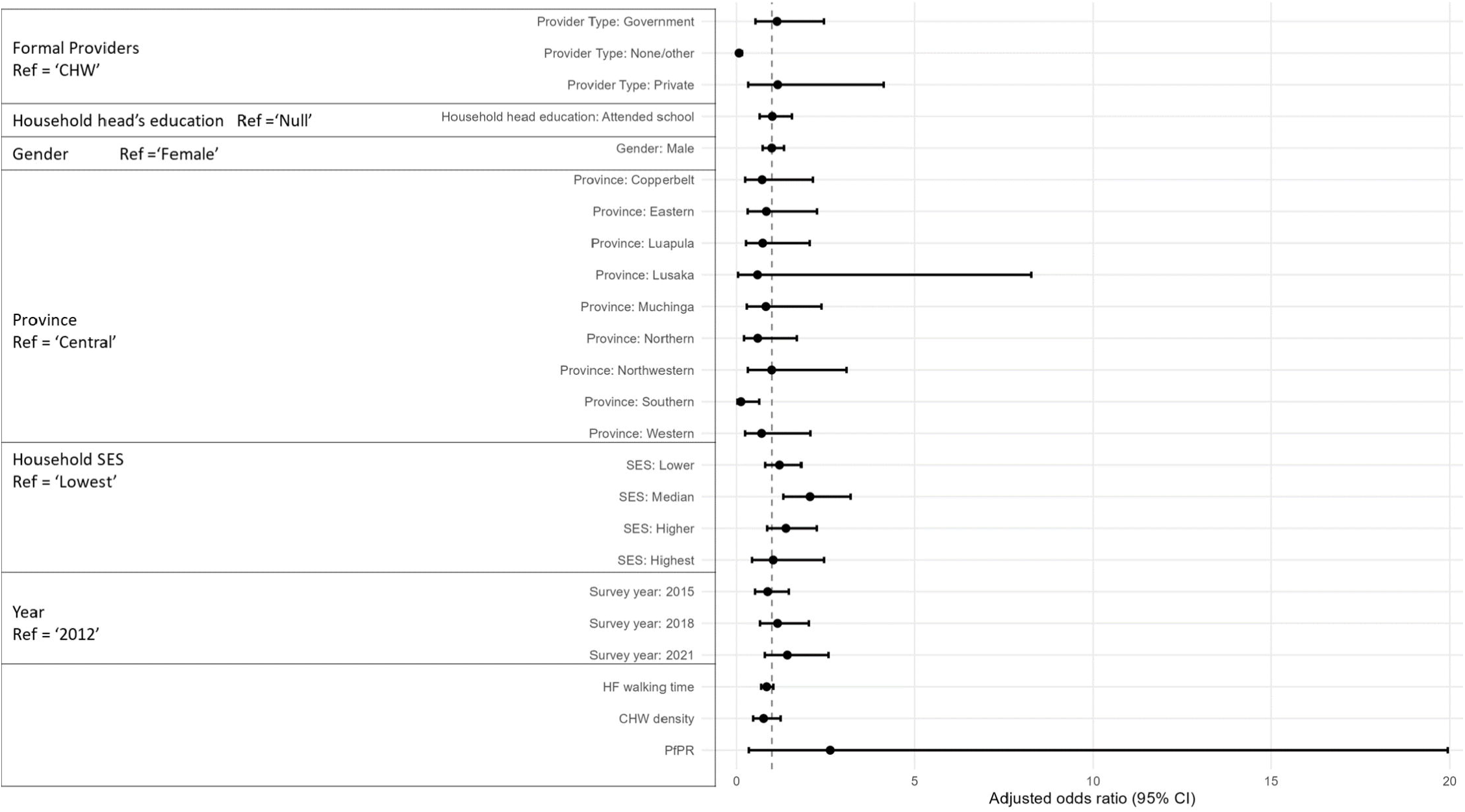
Model 2 outputs. The determinants of AL-receipt in children <5. Results presented are the associated Odds Ratio (OR), 95% confidence interval (CI) from the logit model. N = 1394. Note: Travel time is measured in hours.

## Discussion

In this study, we examined several key aspects of access to malaria diagnosis and treatment in Zambia from 2012 to 2021, in the context of CCM expansion using four nationally representative household surveys. Our investigation focused on whether the scale-up of CCM in Zambia was associated with changes in treatment-seeking for fever over time, and among those who sought treatment, the time between fever onset and care-seeking, as well as the likelihood of seeking care from a CHW. We also assessed the proportion of children < 5 years with malaria who received AL treatment; it is hoped that all children with a fever that have a *Plasmodium* infection should receive the appropriate treatment with AL. Additionally, we explored the determinants of treatment-seeking behavior among febrile individuals and AL receipt among children <5 with malaria.

Malaria provider coverage continuously improved and the walking time to the nearest provider was consistently shortened nationwide. However, there were no significant changes observed in the 4 surveys for the proportion of febrile individuals seeking treatment for a fever nor in the AL receipt among children <5 with malaria between 2012 and 2021, the period in which CCM was scaled-up across Zambia. Substantial increases in malaria provider coverage were identified in Western, Southern, Northwestern, and Eastern Provinces across the period; however, while their treatment-seeking rates maintained at relatively high levels, we did not observe corresponding increases in treatment-seeking behaviors during this period. By contrast, provinces with higher malaria burden such as Luapula, Muchinga, and Northern Provinces experienced both limited increases in malaria provider coverage and consistently low treatment-seeking proportions across time. Likewise, low and temporally fluctuating AL receipt rates were observed across provinces regardless of their provider coverage levels. Of note, by 2021, the average walking time to the nearest provider in all provinces except Muchinga was less than 1 hour, achieving the target goal set by NCHS 2019-2021.(14) The improved walking time coupled with unchanged treatment-seeking rates suggests that interventions beyond reducing physical access to providers are needed to improve febrile patients’ treatment-seeking behavior in Zambia.

Despite these challenges, our study highlighted two notable trends: an increase in the proportion of individuals seeking treatment within 24 hours of fever onset, and a temporary increase in the proportion of formal care seekers reporting care from a CHW between 2012 and 2018. CHW utilization subsequently declined in 2021, indicating that the earlier increase was not sustained despite continued expansion in CHW density. Overall treatment-seeking rates remained stable during CHW expansion, but the composition of treatment-seeking sources shifted modestly toward CHWs and away from informal and private providers. As the proportion of not seeking treatment changed little, this pattern is suggesting redistribution across provider types rather than increased treatment seeking among previously untreated individuals. Nevertheless, a recurring temporal pattern was observed in several provinces. Periods of increasing CHW density in Northwestern, Eastern, Muchinga, Western, and Southern provinces coincided with increases in reported CHW utilization, although these increases in utilization were not sustained in subsequent surveys. A similar pattern was reported by Seidenberg and colleagues (2012), who observed increased use of CHWs among mothers seeking care for children with fever in Mazabuka and Siavonga districts of Southern Province following the implementation of integrated CCM.(31) They attributed the change in treatment-seeking behavior to residents’ enhanced confidence in CHWs’ medical supplies, skills, and knowledge.(31) Furthermore, their study showed that overall treatment-seeking rates did not increase significantly,(31) providing additional evidence that patients shifted between providers. Lastly, the observed increase in the proportion of individuals seeking treatment within 24 hours of fever onset aligned well with the objectives outlined in Zambia’s Malaria Elimination Strategic Plan 2022-2026, which emphasized the importance of early diagnosis and prompt treatment.(13) Notably, although CHW utilization increased through 2018 before declining in 2021, both CHW utilization and overall treatment-seeking rates remained substantially lower than levels reported in several CCM pilot studies from other SSA settings.(32–36) In addition, reductions in health facility utilization reported in some pilot studies were not observed in our analysis.(32) These discrepancies likely reflect differences in study context and design: our estimates are derived from routine, nationally representative household survey data intended to capture real-world treatment-seeking behavior, whereas many CCM pilot studies were conducted in more controlled settings with intensive implementation support and closer supervision. Similar gaps between pilot-study estimates and routine survey–based estimates of CHW utilization have been documented elsewhere, including in Burkina Faso, where routine household surveys reported substantially lower CHW use than contemporaneous CCM pilot evaluations.(36)

Key predictors of treatment-seeking behavior post-fever onset included travel time to health facilities, age, household SES, and province. The statistical significance of travel time aligns with previous findings suggesting CCM deployment promotes the timely diagnosis and treatment for febrile individuals.(6) Moreover, we found that the relationship between travel time and treatment seeking is non-linear, which is consistent with findings from previous studies on travel distance and healthcare utilization.(37–39) Similar to those studies,(38,39) our model 1 also showed that the slope of the decreasing treatment-seeking probability became flatter as travel time increased. This may be explained by the diminishing effect of proximity on treatment-seeking behavior for those living very far from healthcare facilities. The non-significant association between CHW density and treatment seeking also suggests a redistribution of provider utilization rather than an expansion of overall treatment coverage we discussed above. As for AL receipt, children with malaria who did not obtain care from formal providers were less likely to receive AL treatment compared to children taken to CHWs. However, there is no difference in AL-receipt likelihood among different types of formal providers.

There were several limitations existing in our study. First, the definition of malaria case we adopted in this analysis might not accurately reflect the recent malaria history of these young children, due to the time discrepancy between the history of recent fever within the past 2 weeks and the positive RDT result at the time of the survey, as suggested by Bennett and colleagues who initially developed this outcome measure for malaria from household surveys.(23) Similarly, the low sensitivity of RDTs in field settings may lead to some false-negative diagnoses of malaria case.(23,40) Second, the absence of time-referenced data for operational health facilities in Zambia led to the assumption that all facilities remained operational throughout 2012-2021. This may underestimate the travel time to the nearest providers, as respondents could be linked to nearest but non-operating health facilities. Similarly, the definition of ‘active CHW’ used in the study may bias the estimated travel time upward. As malaria case reporting represents only a fraction of the responsibilities undertaken by CHWs, determining the activity status of CHWs solely based on malaria case reporting may overlook a significant proportion of CHWs who are engaged in other critical health-related activities. However, it is acknowledged that CHWs participating in non-diagnosis and treatment activities are less likely to reduce the treatment provider barriers existing in Zambia. Third, due to the fact that tracing each respondent’s treatment-seeking course to specific health facilities or CHWs is not feasible using the MIS, our study omitted the situation where febrile individuals bypassed nearest facility/CHW and went to one further away for whatever reason. Nonetheless, healthcare bypass in the SSA region was found to be prevalent by previous studies.(41–43) Real or perceived stockouts of drugs, lack of health workers, closed health facilities, and poor services provided by nearest clinicians were found able to alter treatment-seeking decision among residents.(41,42) Consequently, the low treatment-seeking depicted in our maps may not solely reflect a shortfall in CCM coverage. They could also indicate broader health system challenges affecting functioning of local health facilities and CHWs.

## Conclusion

Despite the nationwide scale-up of community case management (CCM) over the last decade, significant barriers persist that limit malaria patients’ access to timely diagnosis and effective treatment in Zambia. Our analysis indicates that CCM expansion occurred alongside some improvements, but was not sufficient to address all access challenges. The findings suggest that maintaining and further expanding CCM coverage is important, but must be complemented by additional targeted interventions that address the multifaceted barriers to care.

## Data Availability Statement

The public geospatial datasets used in the analysis are available through the sources cited in the manuscript. The DHIS2 malaria-provider data and Zambia Malaria Indicator Survey microdata were obtained from the National Malaria Elimination Centre and are not publicly redistributable by the authors. Qualified researchers may request access from the Zambia Ministry of Health at or through the National Malaria Elimination Centre at. The authors accessed these data under applicable data-use agreements and had no special access privileges.

## Funding

Financial support for this study was provided by the Bill & Melinda Gates Foundation through a grant awarded to PATH’s Malaria Control and Elimination Partnership in Africa (MACEPA 4 Program, Grant Number: INV-0003212). This grant supported the work of ZM and TE. The funders had no role in the study design, data collection and analysis, decision to publish, or preparation of the manuscript.

## Supporting information

**S1 Table**. Characteristics of individual treatment-seeking behavior in Zambia, 2012–2021. Survey data from 2012, 2015, 2018, and 2021 were re-weighted.

**S2 Table**. Characteristics of individuals receiving artemether–lumefantrine (AL) treatment in Zambia, 2012–2021. Survey data from 2012, 2015, 2018, and 2021 were re-weighted.

**S3 Table**. Proportion of febrile individuals by type of healthcare provider in Zambia, 2012–2021. Individuals who did not seek treatment or who sought treatment from informal providers were included.

**S1 Fig**. Proportion of individuals (all ages) with fever who sought treatment from a formal provider across Zambian provinces, 2012–2021. Formal malaria care providers include public providers, private providers, and community health workers (CHWs). Base map source: GRID3, “ZMB – Operational Districts” dataset (https://data.grid3.org/datasets/zmb-operational-districts/explore), licensed under CC BY 4.0.

**S2 Fig**. Proportion of individuals seeking care from community health workers (CHWs) over time among those who sought treatment from a formal provider across Zambian provinces, 2012–2021. Formal malaria care providers include public providers, private providers, and CHWs. Base map source: GRID3, “ZMB – Operational Districts” dataset (https://data.grid3.org/datasets/zmb-operational-districts/explore), licensed under CC BY 4.0.

**S3 Fig**. Distribution of time between fever onset and treatment-seeking at a formal provider across Zambian provinces, 2012–2021. Formal malaria care providers include both health facilities and CHWs. Base map source: GRID3, “ZMB – Operational Districts” dataset (https://data.grid3.org/datasets/zmb-operational-districts/explore), licensed under CC BY 4.0.

**S4 Fig**. Proportion of children under five years of age with malaria infection who received artemether–lumefantrine (AL) treatment across Zambian provinces, 2012–2021. Data were missing for Southern and Lusaka Provinces where no children tested positive by rapid diagnostic test (RDT) and reported fever in the two weeks preceding the survey. Base map source: GRID3, “ZMB – Operational Districts” dataset (https://data.grid3.org/datasets/zmb-operational-districts/explore), licensed under CC BY 4.0.

**S5 Fig**. Average walking time (minutes) to the nearest malaria care provider by province in Zambia, 2012–2021. Malaria care providers in this analysis include only CHWs and public providers due to the unavailability of comprehensive private provider geolocation data. Base map source: GRID3, “ZMB – Operational Districts” dataset (https://data.grid3.org/datasets/zmb-operational-districts/explore), licensed under CC BY 4.0.

## Notes

### Competing Interest Statement

The authors have declared no competing interest.

### Author Declarations

The public geospatial datasets used in the analysis are available through the sources cited in the manuscript. The DHIS2 malaria-provider data and Zambia Malaria Indicator Survey microdata were obtained from the National Malaria Elimination Centre and are not publicly redistributable by the authors. Qualified researchers may request access from the Zambia Ministry of Health at or through the National Malaria Elimination Centre at. The authors accessed these data under applicable data-use agreements and had no special access privileges This study utilized data exclusively from the Zambia Malaria Indicator Surveys (MIS), conducting secondary analysis of these de-identified datasets. No primary data collection was performed. The datasets were accessed for research purposes on [06/01/2023]. The authors did not have access to any information that could identify individual participants during or after data collection. In accordance with standard research practices for secondary analyses of anonymous public health surveillance data, formal ethics approval was not required for this study.

## Reference

1. World malaria report 2022 [Internet]. [cited 2023 Jul 31]. Available from: https://www.who.int/teams/global-malaria-programme/reports/world-malaria-report-2022

2. Chizema-Kawesha E, Miller JM, Steketee RW, Mukonka VM, Mukuka C, Mohamed AD, et al. Scaling Up Malaria Control in Zambia: Progress and Impact 2005–2008. Am J Trop Med Hyg. 2010 Sep 7;83(3):480–8. doi:10.4269/ajtmh.2010.10-0035

3. Eisele TP, Larsen DA, Walker N, Cibulskis RE, Yukich JO, Zikusooka CM, et al. Estimates of child deaths prevented from malaria prevention scale-up in Africa 2001-2010. Malar J. 2012 Mar 28;11(1):93. doi:10.1186/1475-2875-11-93

4. Mulenga M, Van geertruyden JP, Mwananyanda L, Chalwe V, Moerman F, Chilengi R, et al. Safety and efficacy of lumefantrine-artemether (Coartem®) for the treatment of uncomplicated Plasmodium falciparum malaria in Zambian adults. Malar J. 2006 Aug 21;5:73. doi:10.1186/1475-2875-5-73 PubMed PMID: 16923176; PubMed Central PMCID: PMC1579224.

5. Hamer DH, Ndhlovu M, Zurovac D, Fox M, Yeboah-Antwi K, Chanda P, et al. Improved diagnostic testing and malaria treatment practices in Zambia. JAMA J Am Med Assoc. 2007 May 23;297(20):2227–31. doi:10.1001/jama.297.20.2227 PubMed PMID: 17519412; PubMed Central PMCID: PMC2674546.

6. Win Han Oo, Gold L, Moore K, Agius PA, Fowkes FJI. The impact of community-delivered models of malaria control and elimination: a systematic review. Malar J. 2019 Aug 6;18(1):269. doi:10.1186/s12936-019-2900-1

7. Fong RM, Kaiser JL, Ngoma T, Vian T, Bwalya M, Sakanga VR, et al. Barriers and facilitators to facility-based delivery in rural Zambia: a qualitative study of women’s perceptions after implementation of an improved maternity waiting homes intervention. BMJ Open. 2022 Jul 25;12(7):e058512. doi:10.1136/bmjopen-2021-058512 PubMed PMID: 35879007; PubMed Central PMCID: PMC9328096.

8. Rudasingwa M, De Allegri M, Mphuka C, Chansa C, Yeboah E, Bonnet E, et al. Universal health coverage and the poor: to what extent are health financing policies making a difference? Evidence from a benefit incidence analysis in Zambia. BMC Public Health. 2022 Aug 13;22:1546. doi:10.1186/s12889-022-13923-1 PubMed PMID: 35964020; PubMed Central PMCID: PMC9375934.

9. Institutionalizing integrated community case management (iCCM) to end preventable child deaths [Internet]. [cited 2024 Feb 1]. Available from: https://www.who.int/publications-detail-redirect/9789240006935

10. Integrated community case management in Zambia: Impact Brief [Internet]. [cited 2023 Nov 15]. Available from: https://www.path.org/resources/integrated-community-case-management-zambia-impact-brief/

11. Linn NYY, Kathirvel S, Das M, Thapa B, Rahman MdM, Maung TM, et al. Are village health volunteers as good as basic health staffs in providing malaria care? A country wide analysis from Myanmar, 2015. Malar J. 2018 Jun 20;17(1):242. doi:10.1186/s12936-018-2384-4

12. Brenner JL, Kabakyenga J, Kyomuhangi T, Wotton KA, Pim C, Ntaro M, et al. Can Volunteer Community Health Workers Decrease Child Morbidity and Mortality in Southwestern Uganda? An Impact Evaluation. PLOS ONE. 2011 Dec 14;6(12):e27997. doi:10.1371/journal.pone.0027997

13. National Malaria Elimination Strategic Plan 2022-2026 [Internet]. [cited 2024 Mar 29]. Available from: https://static1.squarespace.com/static/58d002f017bffcf99fe21889/t/632a4cb0fcd87c13d0165372/1663716530614/ZNMESP+2022+to+2026_SIGNED+120722.pdf.

14. CHW Central [Internet]. 2021 [cited 2023 Nov 13]. Zambia National Community Health Strategy, 2019–2021. Available from: https://chwcentral.org/resources/zambia-national-community-health-strategy-2019-2021/

15. Ashton RA, Hamainza B, Lungu C, Rutagwera MRI, Porter T, Bennett A, et al. Effectiveness of community case management of malaria on severe malaria and inpatient malaria deaths in Zambia: a dose–response study using routine health information system data. Malar J. 2023 Mar 17;22(1):96. doi:10.1186/s12936-023-04525-2

16. Zulu JM, Kinsman J, Michelo C, Hurtig AK. Developing the national community health assistant strategy in Zambia: a policy analysis. Health Res Policy Syst. 2013 Jul 20;11(1):24. doi:10.1186/1478-4505-11-24

17. Chanda P, Hamainza B, Moonga HB, Chalwe V, Pagnoni F. Community case management of malaria using ACT and RDT in two districts in Zambia: achieving high adherence to test results using community health workers. Malar J. 2011 Jun 9;10(1):158. doi:10.1186/1475-2875-10-158

18. Weiss DJ, Nelson A, Gibson HS, Temperley W, Peedell S, Lieber A, et al. A global map of travel time to cities to assess inequalities in accessibility in 2015. Nature. 2018 Jan;553(7688):333–6. doi:10.1038/nature25181

19. MAP [Internet]. [cited 2023 Jul 31]. Malaria Atlas Project | Home. Available from: https://malariaatlas.org/

20. Funk C, Peterson P, Landsfeld M, Pedreros D, Verdin J, Shukla S, et al. The climate hazards infrared precipitation with stations—a new environmental record for monitoring extremes. Sci Data. 2015 Dec 8;2(1):1. doi:10.1038/sdata.2015.66

21. WorldPop [Internet]. [cited 2023 May 28]. Open Spatial Demographic Data and Research. Available from: https://www.worldpop.org/

22. Center For International Earth Science Information Network-CIESIN-Columbia University. Global Gridded Relative Deprivation Index (GRDI), Version 1 [Internet]. Palisades, NY: NASA Socioeconomic Data and Applications Center (SEDAC); 2022 [cited 2023 Nov 8]. Available from: https://sedac.ciesin.columbia.edu/data/set/povmap-grdi-v1 doi:10.7927/3XXE-AP97

23. Bennett A, Bisanzio D, Yukich JO, Mappin B, Fergus CA, Lynch M, et al. Population coverage of artemisinin-based combination treatment in children younger than 5 years with fever and Plasmodium falciparum infection in Africa, 2003–2015: a modelling study using data from national surveys. Lancet Glob Health. 2017 Apr 1;5(4):e418–27. doi:10.1016/S2214-109X(17)30076-1 PubMed PMID: 28288746.

24. Swarthout TD, Counihan H, Senga RKK, van den Broek I. Paracheck-Pf accuracy and recently treated Plasmodium falciparum infections: is there a risk of over-diagnosis? Malar J. 2007 May 16;6:58. doi:10.1186/1475-2875-6-58 PubMed PMID: 17506881; PubMed Central PMCID: PMC1890550.

25. Baiden F, Webster J, Tivura M, Delimini R, Berko Y, Amenga-Etego S, et al. Accuracy of rapid tests for malaria and treatment outcomes for malaria and non-malaria cases among under-five children in rural Ghana. PloS One. 2012;7(4):e34073. doi:10.1371/journal.pone.0034073 PubMed PMID: 22514617; PubMed Central PMCID: PMC3325982.

26. Keating J, Miller JM, Bennett A, Moonga HB, Eisele TP. Plasmodium falciparum parasite infection prevalence from a household survey in Zambia using microscopy and a rapid diagnostic test: implications for monitoring and evaluation. Acta Trop. 2009 Dec;112(3):277–82. doi:10.1016/j.actatropica.2009.08.011 PubMed PMID: 19682968.

27. R: The R Project for Statistical Computing [Internet]. [cited 2023 Nov 26]. Available from: https://www.r-project.org/

28. Brooks M, Bolker B, Kristensen K, Maechler M, Magnusson A, McGillycuddy M, et al. glmmTMB: Generalized Linear Mixed Models using Template Model Builder [Internet]. 2023 [cited 2023 Nov 26]. Available from: https://cran.r-project.org/web/packages/glmmTMB/index.html

29. Voeten CC. buildmer: Stepwise Elimination and Term Reordering for Mixed-Effects Regression [Internet]. 2023 [cited 2024 Jan 24]. Available from: https://cran.r-project.org/web/packages/buildmer/index.html

30. Lüdecke (@strengejacke) D, Makowski (@Dom_Makowski) D, Patil (@patilindrajeets) I, Ben-Shachar (@mattansb) MS, Wiernik BM, Waggoner P, et al. see: Model Visualisation Toolbox for “easystats” and “ggplot2” [Internet]. 2023 [cited 2024 Jan 24]. Available from: https://cran.r-project.org/web/packages/see/index.html

31. Seidenberg PD, Hamer DH, Iyer H, Pilingana P, Siazeele K, Hamainza B, et al. Impact of Integrated Community Case Management on Health-Seeking Behavior in Rural Zambia. Am J Trop Med Hyg. 2012 Nov 7;87(5 Suppl):105–10. doi:10.4269/ajtmh.2012.11-0799 PubMed PMID: 23136285; PubMed Central PMCID: PMC3748509.

32. Tiono AB, Kaboré Y, Traoré A, Convelbo N, Pagnoni F, Sirima SB. Implementation of Home based management of malaria in children reduces the work load for peripheral health facilities in a rural district of Burkina Faso. Malar J. 2008 Oct 3;7:201. doi:10.1186/1475-2875-7-201 PubMed PMID: 18834504; PubMed Central PMCID: PMC2570683.

33. Akweongo P, Agyei-Baffour P, Sudhakar M, Simwaka BN, Konaté AT, Adongo PB, et al. Feasibility and acceptability of ACT for the community case management of malaria in urban settings in five African sites. Malar J. 2011 Aug 16;10(1):240. doi:10.1186/1475-2875-10-240

34. Kisia J, Nelima F, Otieno DO, Kiilu K, Emmanuel W, Sohani S, et al. Factors associated with utilization of community health workers in improving access to malaria treatment among children in Kenya. Malar J. 2012 Jul 30;11(1):248. doi:10.1186/1475-2875-11-248

35. Littrell M, Moukam LV, Libite R, Youmba JC, Baugh G. Narrowing the treatment gap with equitable access: mid-term outcomes of a community case management program in Cameroon. Health Policy Plan. 2013 Oct 1;28(7):705–16. doi:10.1093/heapol/czs110

36. Druetz T, Ridde V, Kouanda S, Ly A, Diabaté S, Haddad S. Utilization of community health workers for malaria treatment: results from a three-year panel study in the districts of Kaya and Zorgho, Burkina Faso. Malar J. 2015 Feb 13;14(1):71. doi:10.1186/s12936-015-0591-9

37. Myers CK. Measuring the Burden: The Effect of Travel Distance on Abortions and Births. SSRN Electron J. 2021. doi:10.2139/ssrn.3892584

38. Oldenburg CE, Sié A, Ouattara M, Bountogo M, Boudo V, Kouanda I, et al. Distance to primary care facilities and healthcare utilization for preschool children in rural northwestern Burkina Faso: results from a surveillance cohort. BMC Health Serv Res. 2021 Mar 9;21:212. doi:10.1186/s12913-021-06226-5 PubMed PMID: 33750364; PubMed Central PMCID: PMC7941928.

39. Feikin DR, Nguyen LM, Adazu K, Ombok M, Audi A, Slutsker L, et al. The impact of distance of residence from a peripheral health facility on pediatric health utilisation in rural western Kenya. Trop Med Int Health. 2009;14(1):54–61. doi:10.1111/j.1365-3156.2008.02193.x

40. Chiodini PL, Bowers K, Jorgensen P, Barnwell JW, Grady KK, Luchavez J, et al. The heat stability of Plasmodium lactate dehydrogenase-based and histidine-rich protein 2-based malaria rapid diagnostic tests. Trans R Soc Trop Med Hyg. 2007 Apr 1;101(4):331–7. doi:10.1016/j.trstmh.2006.09.007

41. Bell G, Macarayan EK, Ratcliffe H, Kim JH, Otupiri E, Lipsitz S, et al. Assessment of Bypass of the Nearest Primary Health Care Facility Among Women in Ghana. JAMA Netw Open. 2020 Aug 12;3(8):e2012552. doi:10.1001/jamanetworkopen.2020.12552 PubMed PMID: 32785634; PubMed Central PMCID: PMC7424402.

42. Kahabuka C, Kvåle G, Moland KM, Hinderaker SG. Why caretakers bypass Primary Health Care facilities for child care - a case from rural Tanzania. BMC Health Serv Res. 2011 Nov 17;11(1):315. doi:10.1186/1472-6963-11-315

43. Audo MO, Ferguson A, Njoroge PK. Quality of health care and its effects in the utilisation of maternal and child health services in Kenya. East Afr Med J. 2005 Nov;82(11):547–53. doi:10.4314/eamj.v82i11.9407 PubMed PMID: 16463747.

