## Supplementary material for "Access to malaria diagnosis and treatment in Zambia in the context of scaling-up community case management: results from repeated national cross-sectional surveys": S1_Table.docx

| Indicators | Proportion of febrile individuals seeking treatment | Lower CI | Upper CI | Chi-square statistics |
| --- | --- | --- | --- | --- |
| Province | | | | |
| Central | 0.57 | 0.53 | 0.61 | $X^{2}$= 1732  P-value = 1.712e-10 |
| Copperbelt | 0.66 | 0.60 | 0.71 |  |
| Eastern | 0.70 | 0.66 | 0.74 |  |
| Luapula | 0.47 | 0.43 | 0.51 |  |
| Lusaka | 0.59 | 0.48 | 0.69 |  |
| Muchinga | 0.52 | 0.47 | 0.57 |  |
| North-Western | 0.63 | 0.53 | 0.72 |  |
| Northern | 0.48 | 0.42 | 0.54 |  |
| Southern | 0.57 | 0.51 | 0.63 |  |
| Western | 0.59 | 0.56 | 0.63 |  |
| Year | | | | |
| 2012 | 0.56 | 0.52 | 0.59 |  |
| 2015 | 0.56 | 0.53 | 0.59 |  |
| 2018 | 0.59 | 0.54 | 0.64 | $X^{2}$=26.634  P-value = 0.8292 |
| 2021 | 0.57 | 0.53 | 0.62 |  |
| Gender | | | | |
| Male | 0.57 | 0.55 | 0.60 | $X^{2}$=31.635  P-value = 0.3408 |
| Female | 0.56 | 0.54 | 0.59 |  |
| Urbanicity | | | | |
| Urban | 0.62 | 0.56 | 0.67 | $X^{2}$=165.42  P-value = 0.3192 |
| Rural | 0.56 | 0.53 | 0.57 |  |
| Household socioeconomic status | | | | |
| Lowest | 0.52 | 0.49 | 0.56 | $X^{2}$=439.78  P-value = 0.07858 |
| Lower | 0.52 | 0.49 | 0.56 |  |
| Median | 0.56 | 0.52 | 0.60 |  |
| Higher | 0.60 | 0.56 | 0.63 |  |
| Highest | 0.62 | 0.57 | 0.57 |  |
| Household head’s educational attainment | | | | |
| Yes | 0.57 | 0.55 | 0.59 | $X^{2}$=67.218  P-value = 0.2366 |
| No | 0.54 | 0.50 | 0.59 |  |
| Supplementary table 1. Characteristics of individual treatment seeking behavior 2012-2021. Survey data from 2012, 2015, 2018, and 2021 were re-weighted. | | | | |
