## Supplementary material for "Access to malaria diagnosis and treatment in Zambia in the context of scaling-up community case management: results from repeated national cross-sectional surveys": S2_Table.docx

| Indicators | Proportion of malaria-infected children receiving AL | Lower CI | Upper CI | Chi-square statistics |
| --- | --- | --- | --- | --- |
| Formal providers | | | | |
| CHW | 0.60 | 0.50 | 0.68 | $X^{2}$= 264.58  P-value = 0.1954 |
| Public | 0.66 | 0.61 | 0.71 |  |
| Private | 0.77 | 0.47 | 0.93 |  |
| Year | | | | |
| 2012 | 0.38 | 0.32 | 0.46 | $X^{2}$= 13.999  P-value = 0.8049 |
| 2015 | 0.41 | 0.34 | 0.47 |  |
| 2018 | 0.45 | 0.36 | 0.54 |  |
| 2021 | 0.42 | 0.35 | 0.49 |  |
| Gender | | | | |
| Female | 0.42 | 0.35 | 0.48 | $X^{2}$= 14.687  P-value = 0.5697 |
| Male | 0.41 | 0.35 | 0.46 |  |
| Urbanicity | | | | |
| Urban | 0.45 | 0.27 | 0.65 | $X^{2}$= 3.5354  P-value = 0.7979 |
| Rural | 0.41 | 0.37 | 0.46 |  |
| Household socioeconomic status | | | | |
| Lowest | 0.37 | 0.29 | 0.47 | $X^{2}$= 200.51  P-value = 0.3831 |
| Lower | 0.41 | 0.32 | 0.51 |  |
| Median | 0.52 | 0.36 | 0.68 |  |
| Higher | 0.49 | 0.35 | 0.63 |  |
| Highest | 0.34 | 0.14 | 0.62 |  |
| Supplementary table 2. Characteristics of individual receiving AL 2012-2021. Survey data from 2012, 2015, 2018, and 2021 were re-weighted. | | | | |
