## Supplementary material for "Access to malaria diagnosis and treatment in Zambia in the context of scaling-up community case management: results from repeated national cross-sectional surveys": S3_Table.docx

| MIS year | Public facility | CHW | Private facility | No-formal providers |
| --- | --- | --- | --- | --- |
| 2012 | 43.1 [39.6, 46.7] | 0.7 [0.4, 1.2] | 2.7 [1.5, 4.6] | 53.5 [49.3, 57.7] |
| 2015 | 41.2 [37.5, 44.9] | 3.5 [2.3, 5.4] | 1.5 [0.5, 4.1] | 53.8 [50.1, 57.6] |
| 2018 | 45.4 [39.8, 51.2] | 5.1 [2.8, 9.0] | 0.1 [0.0, 0.3] | 49.4 [43.5, 55.3] |
| 2021 | 50.0 [45.1, 54.9] | 1.7 [0.7, 3.7] | 0.6 [0.3, 1.2] | 47.7 [42.8, 52.7] |
| Supplementary table 3. Proportion of febrile individuals by type of healthcare provider in Zambia, 2012-2021" *Individuals who didn’t seek treatment or sought treatment from an informal provider were included | | | | |
