## Supplementary figures and images for "Access to malaria diagnosis and treatment in Zambia in the context of scaling-up community case management: results from repeated national cross-sectional surveys"

### S1_Fig.tiff

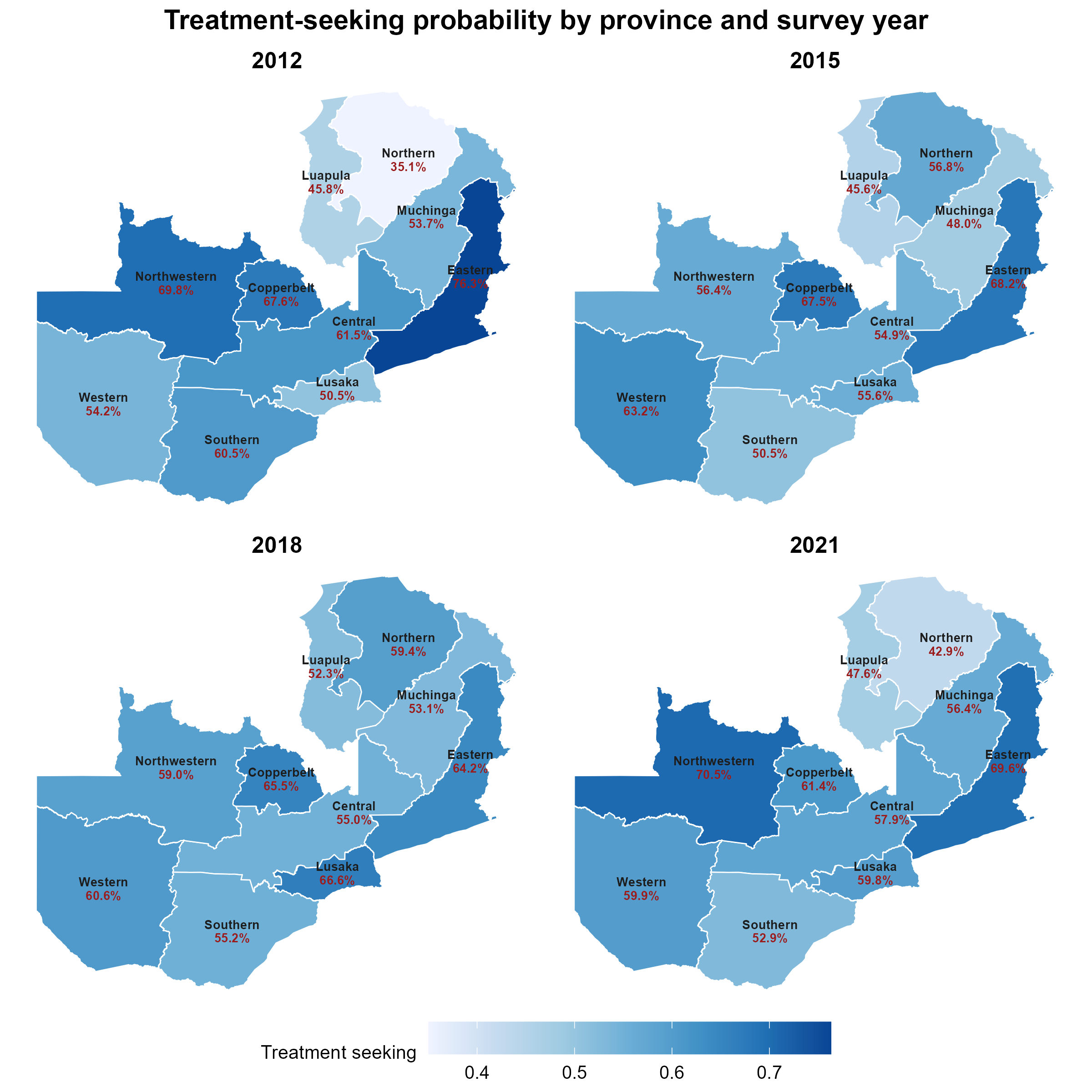

### S2_Fig.tiff

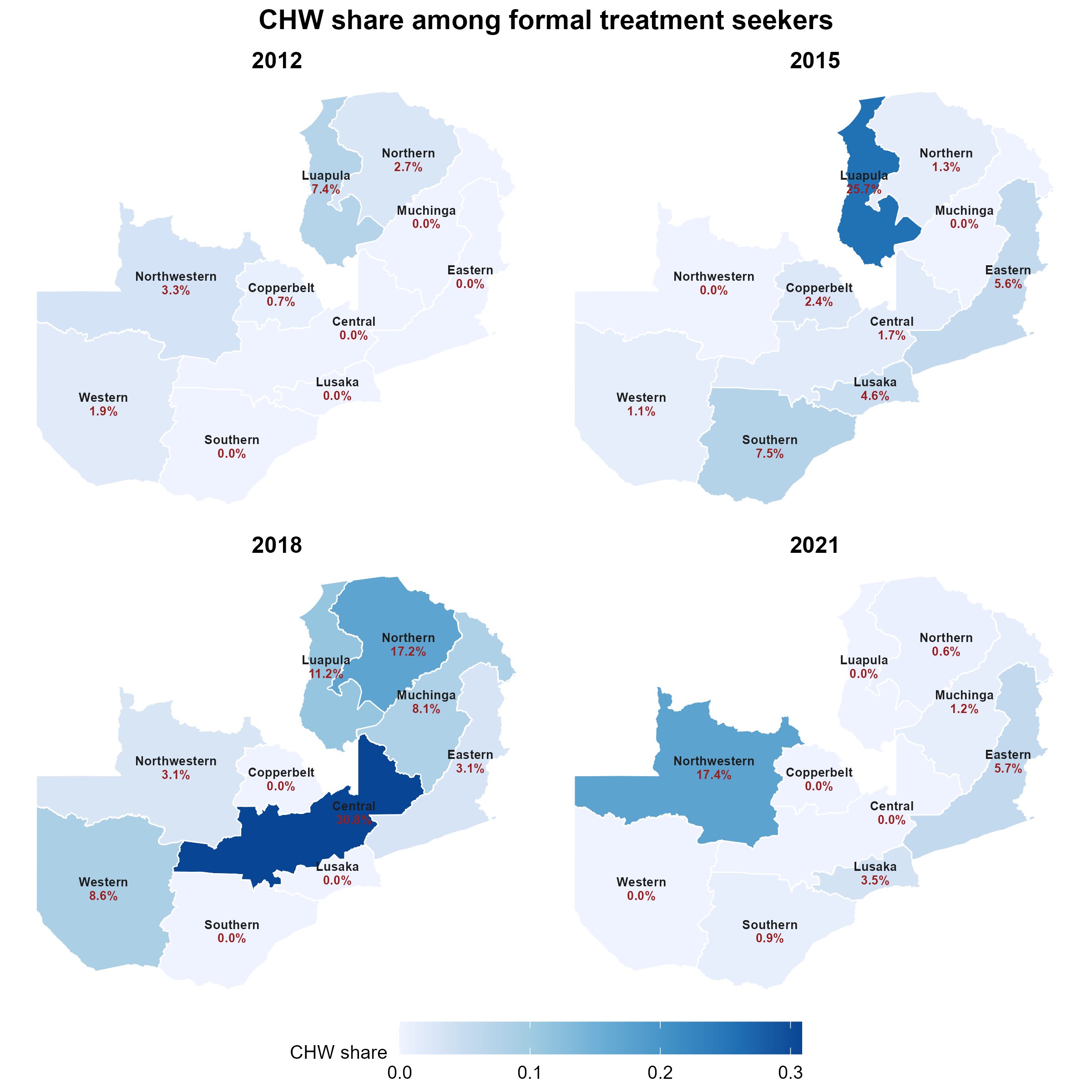

### S3_Fig.tiff

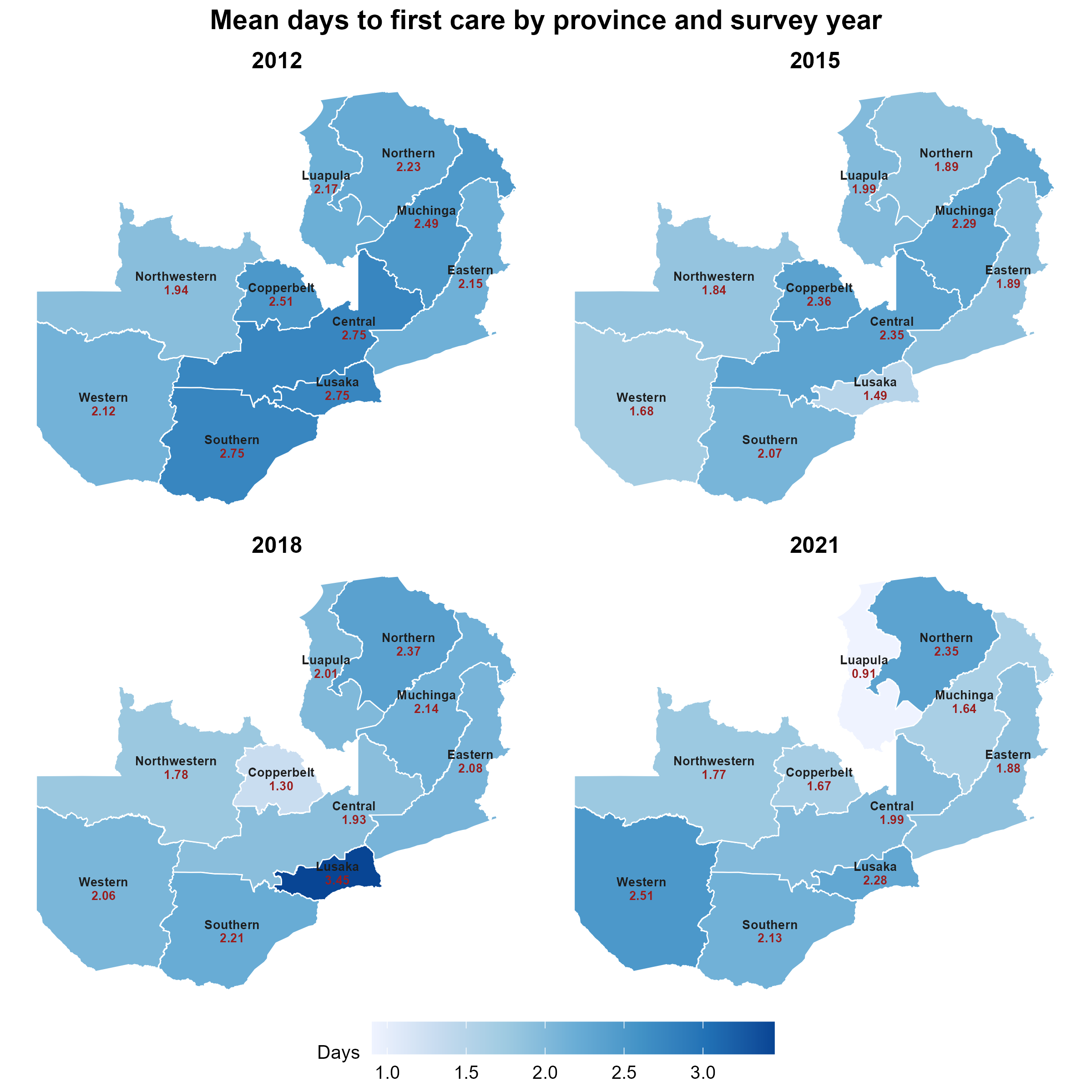

### S4_Fig.tiff

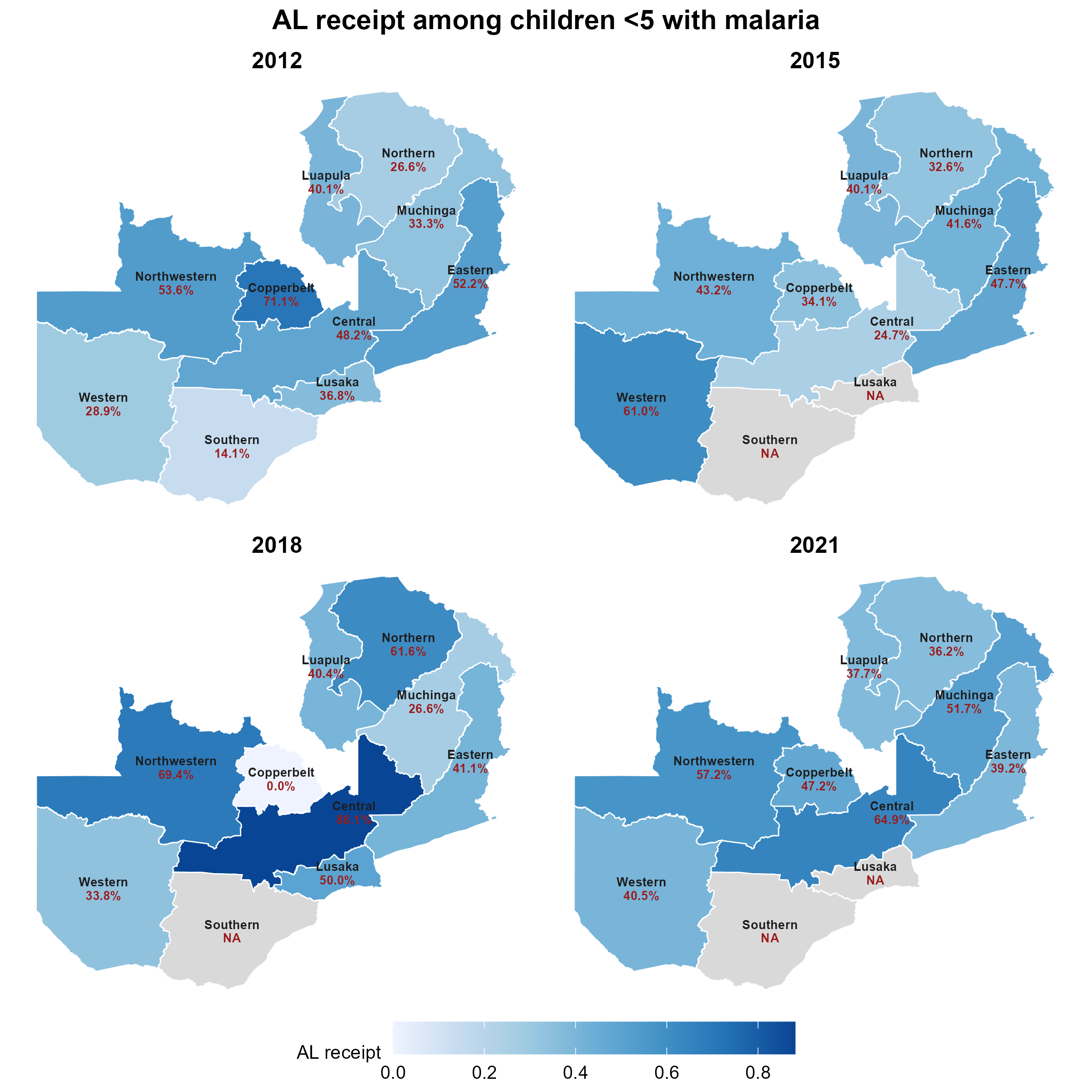

### S5_Fig.tiff

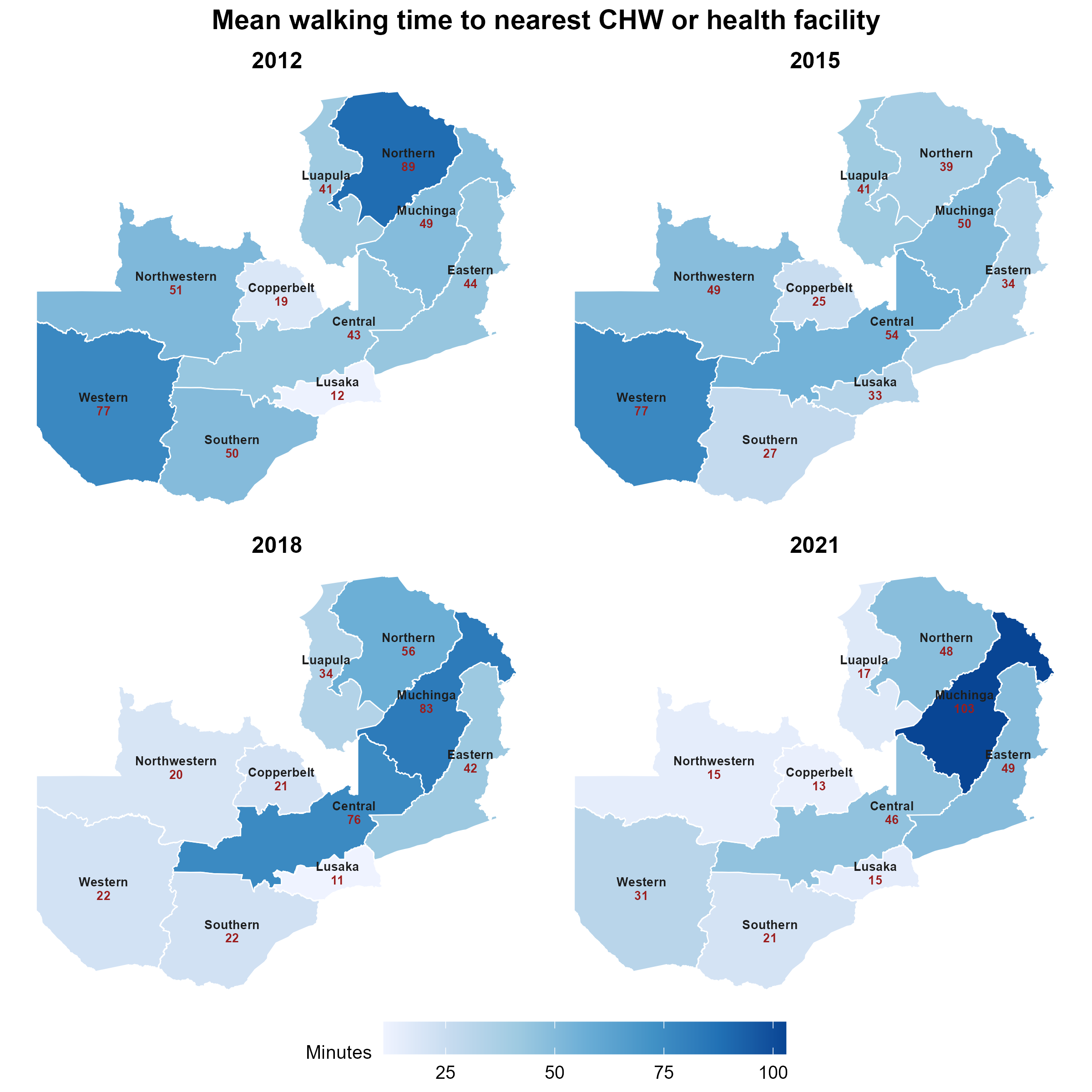
